# Playing-related problems in guitarists: Systematic review and meta-analysis

**DOI:** 10.1101/2025.10.08.25337619

**Authors:** Mario Castillo-Barragán, Laura Leticia Tirado-Gómez, José Gabriel García-Palacios, Jorge Miguel Gutiérrez-Libreros, Juan Rodrigo Gómez-Bernal

## Abstract

**Background:** Playing-related disorders (PRP) are among the most common occupational health problems affecting musicians; however, no prior meta-analysis has focused exclusively on guitarists. Existing reviews often group plucked-string players with bowed-string or other instrumentalists, overlooking the unique biomechanical demands of guitar performance. In addition, the widely used term PRMD (playing-related musculoskeletal disorders) frequently conflates clinically diagnosed conditions with self-reported symptoms, limiting clinical clarity. We propose a classification under the umbrella term playing-related problems (PRP) comprising: (1) PRMS—self-reported playing-related musculoskeletal symptoms; (2) PRMD—clinically diagnosed playing-related musculoskeletal disorders; and (3) PRND—clinically diagnosed playing-related neurological disorders. All are secondary to instrument playing and defined by the presence of a symptom (PRMS) or disorder (PRMD or PRND) that interferes with the musician’s ability to perform at their usual level.

**Methods:** Following PRISMA guidelines, we conducted a systematic review and meta-analysis of PRP prevalence in guitarists. Studies were classified by PRP subtype. Random-effects meta-analyses of proportions were performed, with subgroup analyses and mixed-effects meta-regressions to explore heterogeneity. Funnel plots and Egger’s test were used to assess potential publication bias.

**Results:** Twenty-two cross-sectional studies were included (n = 2,119). Pooled prevalence was 51.8% for any PRP, 64.5% for PRMS, 42.2% for PRMD, and 14.5% for PRND. In analyses restricted to musculoskeletal outcomes (PRMS + PRMD), specifying guitar type substantially reduced heterogeneity, with classical, bass, and flamenco guitarists showing the highest prevalences. Meta-regression identified PRP subtype, guitar type, and prevalence recall period as significant moderators; residual heterogeneity (I²) decreased from 90.7% to 0% in the most restricted model. No evidence of funnel-plot asymmetry was found for PRP. Qualitative synthesis suggested possible associations with daily practice time, warm-up habits, sex, and multi-instrumentalism, although findings were inconsistent.

**Conclusions:** PRP are highly prevalent among guitarists. Adopting the PRP– PRMS–PRMD–PRND framework may improve consistency in case definitions, facilitate valid prevalence comparisons, and support the development of instrument-specific preventive and rehabilitative strategies. Evidence from secondary outcomes suggests that incorporating warm-up routines and practice breaks may offer additional protection against these disorders.

## Introduction

Playing-related musculoskeletal disorders (PRMD) are among the most common occupational health problems affecting both professional and student musicians, with a reported lifetime prevalence ranging from 39% to 87%, depending on the population and definition used [1,2]. These conditions can severely limit practice time, diminish performance quality, and ultimately shorten professional careers [3]. Within this context, guitarists represent a particularly vulnerable population due to the unique and strenuous biomechanical demands of their practice. The traditional seated posture of classical guitar, with the instrument resting on a raised thigh, creates asymmetric spinal loading and prolonged trunk flexion, which are strongly associated with chronic back and shoulder pain. The demands on the hands are equally extreme and distinct: the fretting hand must apply continuous finger pressure and maintain awkward wrist positions, while the plucking hand executes rapid, repetitive movements, often for several hours a day [4].

Although evidence points to a high prevalence of these issues in guitarists [5–7], a consolidated understanding is hindered by several key methodological limitations. One major challenge is the inconsistent and imprecise use of terminology. The term PRMD is often applied broadly, conflating clinically diagnosed musculoskeletal conditions such as tendinitis with any self-reported symptom of pain or discomfort that interferes, to some degree, with musical performance [7–9]. As noted by previous authors, this conceptual ambiguity complicates both research synthesis and clinical interpretation [10]. Furthermore, it is compounded by a lack of specificity regarding guitarist subpopulations, as most studies fail to differentiate between playing different type of guitars (e.g., classical, electric, acoustic) or aggregate them into a single group, introducing substantial heterogeneity and limiting the conclusions [11].

To address these issues, we propose the broader construct Playing-Related Problems (PRP), defined as any playing-associated condition that prevents the guitarist from performing at their previous level. Within this framework, we distinguish three subcategories: (1) Playing-Related Musculoskeletal Symptoms (PRMS) — self-reported musculoskeletal complaints perceived by the musician as limiting or impairing their ability to perform the instrument as they were accustomed to, without a confirmed diagnosis; (2) Playing-Related Musculoskeletal Disorders (PRMD) — musculoskeletal injuries, disorders, or diseases diagnosed by a healthcare professional (e.g., overuse syndrome, tendinitis); and (3) Playing-Related Neurological Disorders (PRND) —clinically diagnosed neurological conditions associated with playing (e.g., focal dystonia, playing-related neuropathies).

This conceptual clarification is central to our analytical approach. Accordingly, we conducted a comprehensive systematic review and meta-analysis of PRPs in guitarists, aiming to generate more accurate and clinically relevant prevalence estimates by explicitly accounting for definitional heterogeneity, stratifying data by problem type, and comparing prevalence between studies of classical guitarists and those with mixed or unspecified guitarist populations.

## Methods

### Protocol and Registration

This systematic review and meta-analysis were conducted and reported following the PRISMA (Preferred Reporting Items for Systematic Reviews and Meta-Analyses) guidelines [12]. Details of compliance are provided in S1 File. The study protocol was pre-registered in the PROSPERO international prospective register of systematic reviews (registration no. CRD420251069560). It should be noted that while the initial protocol title and objectives focused exclusively on classical guitarists, the systematic literature search revealed that most of the available studies did not specify the guitarist type. The review’s scope was broadened to include all guitar types to address this limitation in the evidence and provide a more comprehensive analysis. Consequently, the analysis plan was updated to incorporate the subgroup analyses and the stratification of PRPs as detailed in this manuscript. These amendments were documented in the PROSPERO record before data extraction.

### Eligibility Criteria and PICO Framework

Population was defined as guitarists of any age, with an emphasis, but not exclusivity, on classical guitarists (formally trained in classical technique). We also included studies of a mixed population of guitarists, provided data specific to classical or acoustic guitarists that were available, and regular guitar playing (≥3 months of formal or self-directed practice). In risk-factor analyses, the “exposure” of interest included high daily practice time, posture variables (e.g., seated with a footstool vs. alternative support or standing posture), psychosocial stress, and a history of prior injury. In addition to the prevalence estimations in the meta-analysis, we compared classical versus (1) non-classical guitarists (e.g., acoustic, bass, electric, flamenco) and (2) within neurological vs musculoskeletal problems. The primary outcome was studies that reported any PRP, which included PRMD, defined as conditions requiring health professional evaluation for confirmation; 2) self-reported musculoskeletal symptoms, identified solely through questionnaires or surveys with some grade affecting of the guitar technique (PRMS); and PRND, such as clinical diagnosed by an health expert (such a neuropathies or focal dystonia). We included observational studies (cross-sectional, cohort, and case-control) and clinical studies (e.g., case series) that provided data on prevalence or risk factors. Intervention studies (e.g., trials of preventive exercises) were also considered if they reported baseline prevalence or other relevant observational data, although none were ultimately found that met our criteria. We excluded studies where guitarists were grouped with other instrumentalists, and data for guitarists could not be distinguished from those of other instrumentalists, and systematic or scoping reviews were not considered primary data; however, we mined them for additional references.

### Information sources and search strategy

We conducted a systematic literature search across five electronic databases, PubMed, Scopus, Web of Science, Dimensions, and PsycINFO. The search strategy combined terms related to musicians and musculoskeletal disorders, for example: “(guitar OR guitarist*) AND (musculoskeletal OR injury OR pain OR disorder)”. No date and language restrictions were applied, and the final searches were conducted on June 7, 2025. In addition to database searches, we performed snowball sampling of references using Connected Papers and citation tracking. We also manually searched the journal Medical Problems of Performing Artists (a core journal in performing arts medicine) and reviewed conference proceedings in performing arts health for relevant abstracts, and searches were not limited to language restrictions. The entire strategy for all databases is provided in the S2 File.

### Study Selection and Data Extraction

All citations were imported into Rayyan software, where duplicates were automatically removed and then manually confirmed. Two reviewers [MCB and JRGB] independently screened titles and abstracts for relevance. The full texts of potentially eligible studies were then assessed against the criteria. Any disagreements in inclusion were resolved by a third reviewer [LLTG]. From each included study, we extracted key data, including publication details (year, country), study design, sample size and demographics (age, sex, professional status of guitarists), definition of musculoskeletal or neurological outcome (e.g., any pain vs. only disorders affecting performance), prevalence of PRPs (point, 12-month, or lifetime as reported), and any risk factors or subgroup results (guitar type, musculoskeletal and neurological injury). For studies that included multiple instrument types or a comparison between classical and other guitarists, we extracted the relevant subgroup data (for example, separate prevalence for classical vs. flamenco guitarists in a study that compared them).

### Study Risk of Bias Assessment

We assessed the risk of bias in the included studies using criteria adapted from the Joanna Briggs Institute checklist for prevalence purposes [13]. Key factors considered included the sampling method (random or convenience sample), response rate, case definition of musculoskeletal disorder (and whether it required symptoms to affect play), recall period for prevalence (to assess potential recall bias), and statistical rigor in any risk factor analysis. Overall, most studies were cross-sectional in design, which inherently limits the ability to make causal inferences.

### Data Synthesis and Analysis

All statistical analyses were performed in R (v4.4.1; R Foundation for Statistical Computing, Vienna, Austria) using the “meta” and “metafor” packages. We performed a random-effects meta-analysis of proportions to calculate pooled prevalence estimates and their 95% confidence intervals (CIs) for each predefined category; all PRP, clinically diagnosed PRMD, self-reported PRMS, and clinically diagnosed PRND. The Freeman–Tukey double arcsine transformation was applied to stabilize the variance of proportions. Heterogeneity was assessed using Cochran’s Q test and quantified with the I² statistic, where values >50% were considered indicative of substantial heterogeneity; p-values <0.10 in the Q test were considered statistically significant.

Pre-specified subgroup analyses were conducted where sufficient data were available, including by guitar type and PRP type. To explore potential sources of heterogeneity, we conducted mixed-effects meta-regression analyses. Given that PRP analyses combined musculoskeletal (high prevalence) and neurological (low prevalence) conditions, type of guitar was not included as a moderator in Models 1 and 2 because its explanatory value would differ markedly between these clinical domains (e.g., Flamenco guitar is associated with high prevalence in musculoskeletal outcomes but much lower prevalence for focal dystonia or neuropathy). For musculoskeletal-only (PRMD and PRMS) analyses (models 3–5), type of guitar was included as a moderator along with age group, prevalence reference period, and mean daily practice hours where available. Residual heterogeneity (τ², I²) was reported for all models. Potential publication bias was evaluated by visual inspection of funnel plots and Egger’s regression test. Sensitivity analyses included: (1) restricting to studies with clearly specified guitar type, (2) restricting to age group (e.g., children and adolescent), (3) restricting to studies with clinically confirmed PRMD or PRND, (4) excluding studies with <30 participants and (5) stratifying by the reported prevalence recall period (12-month, lifetime, three-month, and point prevalence). We also qualitatively synthesized common risk factors (e.g., gender, practice habits, posture) and frequently affected anatomical sites or diagnoses, given the heterogeneity in reporting of risk estimates across studies (S4 File). The overall certainty of the evidence for each outcome was assessed using the Grading of Recommendations Assessment, Development and Evaluation (GRADE) approach, considering study design, risk of bias, inconsistency, indirectness, and imprecision.

## Results

### Study Selection and Characteristics

Our research and selection process identified 22 studies [4–7,14–31] that met the inclusion criteria (PRISMA flow diagram, Fig 1). S1 Table presents all characteristics and main findings of the included studies. These studies, published between 1998 and 2022, collectively included 2,119 guitarists, with individual sample sizes ranging from 14 to 520 participants. Most of the studies were cross-sectional surveys of musicians [4–7,16–26,28–31], conducted in academic, festival, or community settings and using self-administered questionnaires to assess PRP. Only a minority incorporated clinical examinations or objective diagnostic tests, such as physician-confirmed diagnoses [4,14,15,29] or nerve conduction studies with a clinical evaluation [19,22,27].

**Fig 1.**
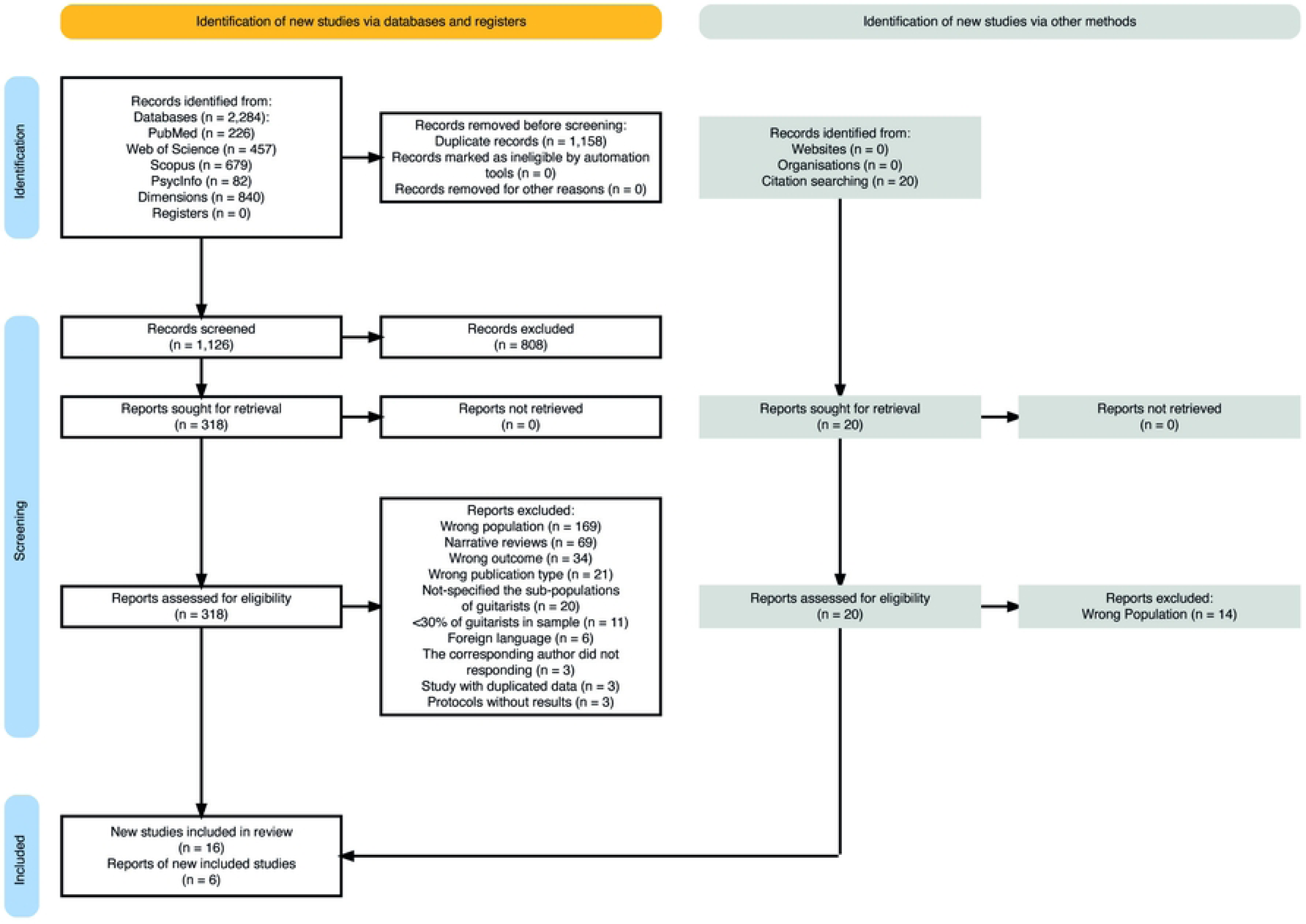
PRISMA flow diagram. PRISMA flowchart depicting the identification, screening, eligibility assessment, and inclusion of studies in the review

Although all studies evaluated PRP attributable to guitar playing, case definitions varied widely. Some considered any pain, discomfort, numbness, or weakness reported as playing-related, whereas others applied stricter definitions, such as symptoms severe enough to interfere with performance or meeting clinical diagnostic criteria (e.g., overuse syndrome, focal dystonia) [4,19,22,24,27,29]. Most studies focused exclusively on guitarists but did not specify the type of guitar; populations included classical [4,17,23,24,26,28], flamenco [4], bass [6,30,31], acoustic or electric guitarists, either exclusively or in mixed groups [7,20,23,25].

Study populations ranged from children and adolescents [17,26] to adults with mean participant ages typically between 18 and 40 years. Both amateur/student and professional/teacher guitarists were represented, although some studies focused on specific groups such as conservatory students [30,31] or festival participants [4,17,28]. Studies were conducted across diverse geographical regions, including North America [5–7,14,15,21,27], Europe [17,19,20,28,30,31], Asia [16,18,23,25,29], Latin America [22,24] and Oceania [26].

### Risk of bias in studies

The body of evidence carries a moderate-to-high risk of bias for prevalence synthesis (Fig 2), yet it still provides valuable insights into symptom patterns among guitarists and related instrumentalists. All included studies represent level 4 evidence in the evidence hierarchy, consisting of cross-sectional designs based on clinical records or survey analyses; no randomized trials or cohort studies were identified in this field. Across the set of studies, every design relied on convenience-based, single-site, or self-selected samples, and none provided denominator data sufficient for population-level prevalence estimates. Most investigations aimed to describe the proportions of musculoskeletal or neurological problems (e.g., pain sites, focal dystonia, neuropathies) in readily accessible musician cohorts, rather than estimating population prevalence. Consequently, external validity is uniformly limited, and uncertainty regarding response rates persists. Nonetheless, internal comparisons, measurement validity, and statistical analyses were generally adequate for their exploratory aims. A detailed risk-of-bias assessment for each study is provided in S1 Fig.

**Fig 2.**
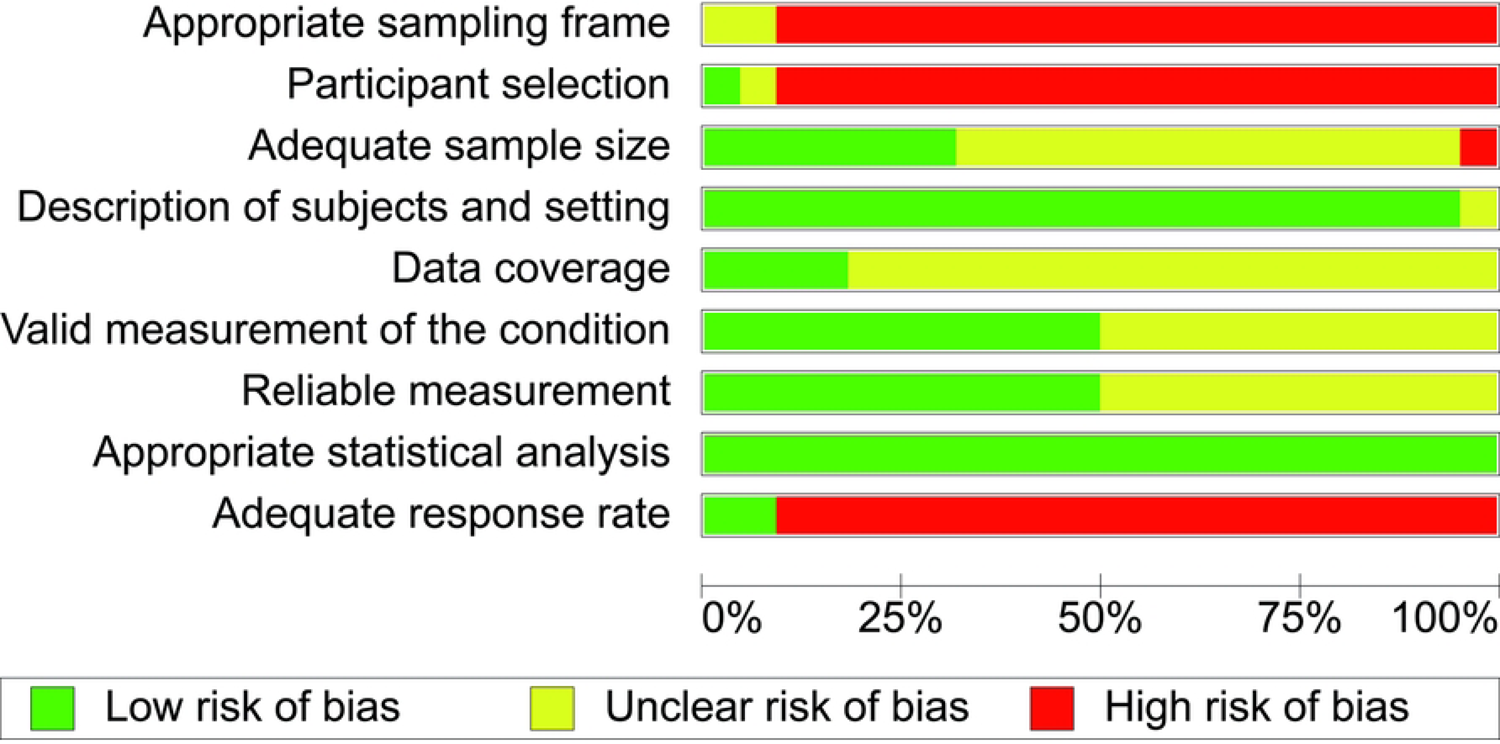
Risk of bias assessment for included studies. Summary of the risk of bias assessment across all included studies based on the Joanna Briggs Institute Critical Appraisal Checklist for Studies Reporting Prevalence Data. Each bar represents the proportion of studies rated as low risk of bias (green), unclear risk of bias (yellow), or high risk of bias (red).

### Prevalence of playing-related problems in guitarists

All studies converged on the finding that musculoskeletal problems are prevalent among guitarists, across 22 studies (n = 2,119 guitarists; 1,201 cases), the pooled prevalence of any PRP was 51.8% (95% CI: 39.2–64.2), with substantial heterogeneity (I² = 95.5%). Subgroup analysis by problem type revealed significant differences (Q = 24.73, p < 0.0001) (Fig 3). The highest prevalence was observed for PRMS (64.5%, 95% CI: 53.9–73.7; I² = 92.0%), followed by PRMD (42.2%, 95% CI: 19.8–68.3; I² = 91.4%). PRND were less common, with a pooled prevalence of 14.5% (95% CI: 6.9–28.1; I² = 92.1%). Heterogeneity remained high within each subgroup, indicating that variability was not fully explained by problem type alone.

**Fig 3.**
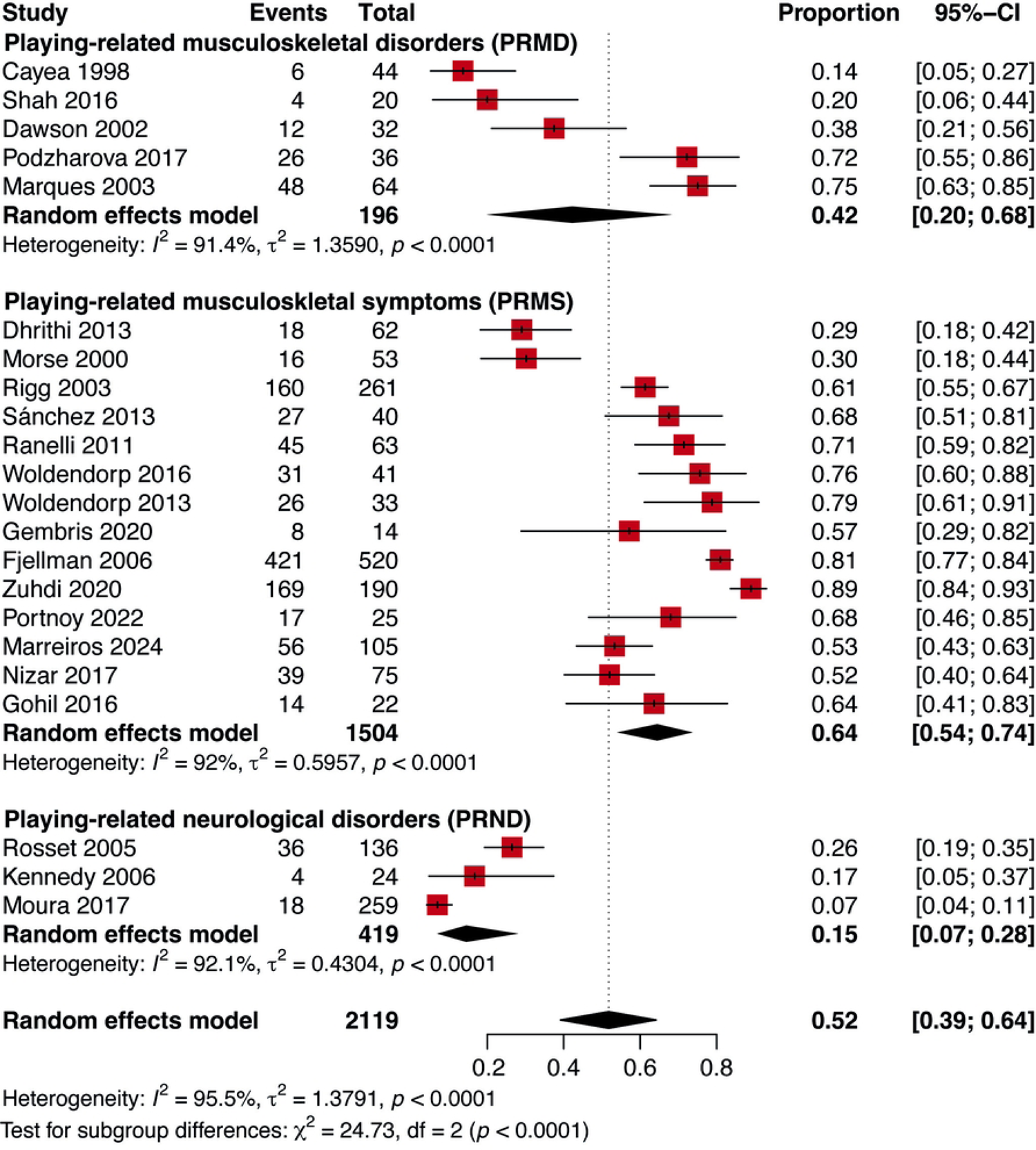
Pooled prevalence of PRP in guitarists by problem type. Random-effects meta-analysis of the pooled prevalence of playing-related problems (PRP) in guitarists, stratified by problem type. Diamonds represent pooled prevalence estimates with 95% confidence intervals (CIs) for each subgroup and for the overall sample. The size of the square markers is proportional to the study weight in the meta-analysis. Heterogeneity statistics (I²) are shown for each subgroup and for the overall estimate.

Across 19 studies reporting musculoskeletal problems PRMD and PRMS (n = 1,301 guitarists; 816 cases), the pooled prevalence was 60.6% (95% CI: 50.3–69.9), with substantial heterogeneity (I² = 88.1%). Subgroup analysis by guitar type revealed significant differences (Q = 28.07, p < 0.0001) (Fig 4). Studies with unspecified guitar type reported the lowest prevalence (40.7%, 95% CI: 20.8–64.3) and the highest heterogeneity (I² = 95.3%). In contrast, studies focusing on Classical guitar (66.9%, 95% CI: 60.3–73.0) and Bass guitar (76.9%, 95% CI: 70.3–82.5) showed no heterogeneity (I² = 0%). The highest prevalence was observed in the single studies of Flamenco guitar (87.5%) and Banjo (78.6%). Mixed guitar types had intermediate prevalence (58.2%, 95% CI: 52.2– 63.9) with moderate heterogeneity (I² = 37.1%). These results indicate that specifying the instrument type substantially reduces heterogeneity and reveals distinct prevalence patterns.

**Fig 4.**
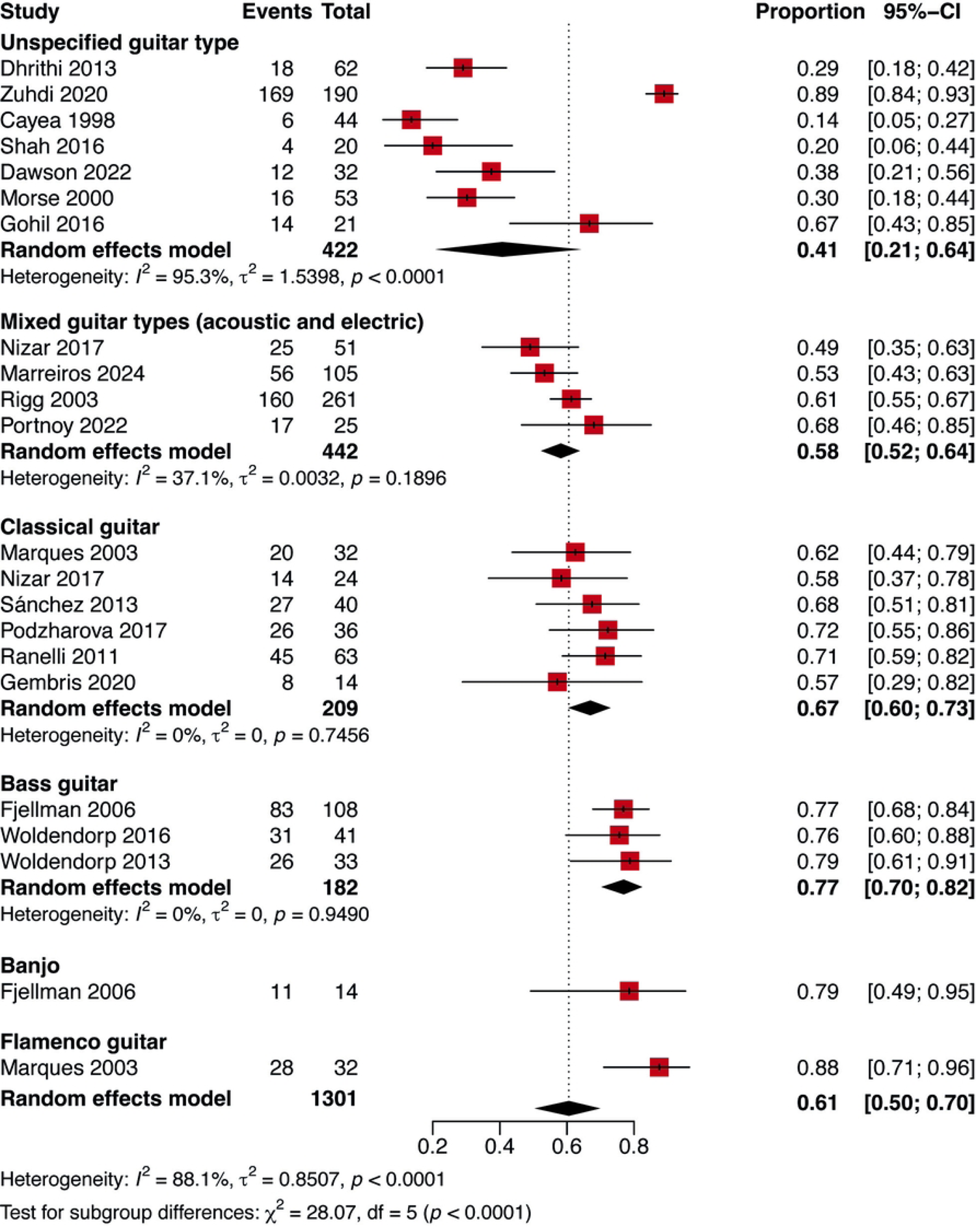
Pooled prevalence of playing-related musculoskeletal problems (PRMD and PRMS) in guitarists by guitar type. Random-effects meta-analysis of the pooled prevalence in guitarists, stratified by guitar type. Diamonds represent pooled prevalence estimates with 95% confidence intervals (CIs) for each subgroup and for the overall sample. The size of the square markers is proportional to the study weight in the meta-analysis. Heterogeneity statistics (I²) are shown for each subgroup and for the overall estimate.

Three studies [14–16] assessed PRND (n = 419 guitarists; 58 cases). The pooled prevalence was 14.5% (95% CI: 6.9–28.1), with substantial heterogeneity (I² = 92.1%). Subgroup analysis by guitar type (Fig 5) showed significant differences (Q = 25.38, p < 0.0001). The highest prevalence was reported in studies including both Classical and Flamenco guitarists (26.5%, 95% CI: 19.7– 34.5), followed by studies mixed Acoustic, Bass, Classical, and Electric guitarists (16.7%, 95% CI: 6.4–36.9) and the lowest prevalence was observed among Acoustic guitar players (7.0%, 95% CI: 4.4–10.8).

**Fig 5.**
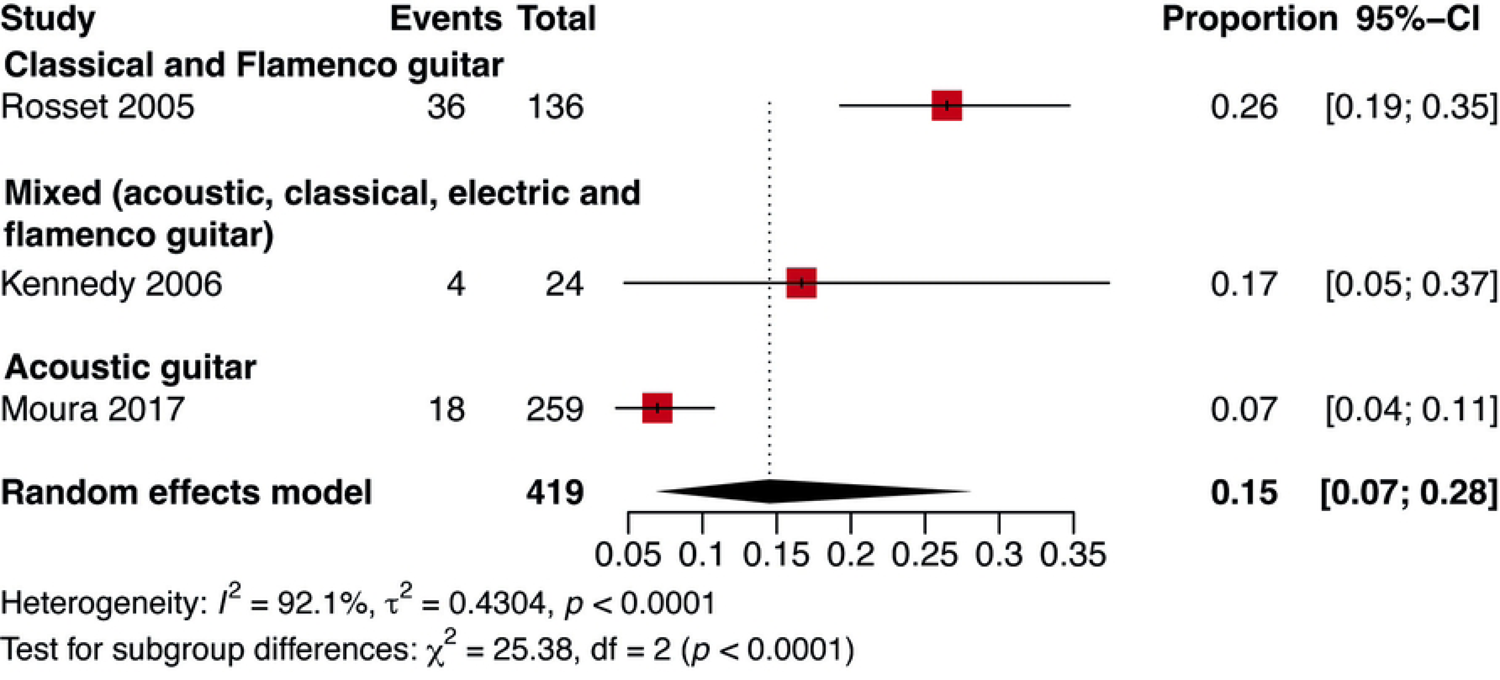
Pooled prevalence of PRND in guitarists. Random-effects meta-analysis of the pooled prevalence in guitarists. Diamonds represent pooled prevalence estimates with 95% confidence intervals (CI) for the overall sample. The size of the square markers is proportional to the study weight in the meta-analysis. Heterogeneity statistics (I²) are shown for the overall estimate.

### Heterogeneity, Sensitivity, and Evidence Quality

To further explore heterogeneity, we conducted mixed-effects meta-regression models (Table 1). In the full PRP models, problem type was a strong predictor: compared to PRND, both PRMD and PRMS were associated with significantly higher prevalence estimates (Model 1: β = 1.45, p = 0.037; β = 2.29, p = 0.0002, respectively). In musculoskeletal-only analyses, guitar type was an important determinant, with Classical, Bass, Flamenco, and Banjo all showing higher prevalence than Unspecified guitar type (Models 3–5; all p < 0.05).

**Table 1.** Summary of meta-regression models evaluating moderators of prevalence estimates for PRP in guitarists.

| Moderator | PRP |  | MSK (PRMD and PRMS) |  |  |
| --- | --- | --- | --- | --- | --- |
|  | Model 1<br>(n=22) | Model 2<br>(n=12) | Model 3<br>(n=19) | Model 4<br>(n=19) | Model 5<br>(n=9) |
| **Problem type (ref: PRND)** |  |  |  |  |  |
| PRMD | 1.41 (0.08, 2.74)* | 2.94 (1.16, 4.73)* | Reference | Reference | — |
| PRMS | 2.25 (1.00, 3.50)* | 2.16 (0.86, 3.47)* | 0.89 (0.34, 1.72)* | 0.89 (0.06 to 1.71)* | — |
| **Guitar type (ref: Unspecified)** |  |  |  |  |  |
| Banjo | — | — | 2.00 (0.09, 3.90)* | 2.33 (0.47, 4.29)* | 1.63 (0.27, 2.99)* |
| Bass guitar | — | — | 1.87 (0.84, 2.91)* | 2.19 (0.79, 3.59)* | 1.95 (1.26, 2.64)* |
| Classical guitar | — | — | 1.01 (0.08, 1.94)* | 1.33 (0.42, 2.23)* | 3.43 (2.10, 4.76)* |
| Flamenco guitar | — | — | 2.65 (0.90, 4.40)* | 2.98 (1.28, 4.68)* | 4.61 (3.12, 6.10)* |
| Mixed guitar types | — | — | 0.57 (-0.29, 1.43) | 0.57 (-0.27, 1.42) | **2.40 (1.06, 3.73)\*** |
| **Age group (ref: Children)** |  |  |  |  |  |
| Middle aged | -0.29 (-2.82, 2.25) | -0.37 (-2.73, 1.98) | -0.18 (-2.13, 1.76) | -1.22 (-3.18, 0.74) | **-2.52 (-3.73, -1.30)\*** |
| Mixed | -0.38 (-2.21, 1.45) | -0.14 (-1.99, 1.70) | 0.13 (-1.50, 1.76) | -0.27 (-1.79, 1.24) | **1.61 (0.59, 2.64)\*** |
| Young adults | -0.46 (-2.24, 1.32) | -0.65 (-2.61, 1.31) | -0.56 (-2.11, 0.99) | -0.61 (-2.02, 0.81) | **-0.90 (-1.77, -0.03)\*** |
| Daily practice (hours/day) | — | -0.07 (-0.39, 0.26) | — | — | **-0.83 (-1.14, -0.53)\*** |
| **Prevalence reference period (ref: shorter)** |  |  |  |  |  |
| Lifetime | -0.10 (-1.25, 1.04) | — | — | **-1.03 (-1.93, -0.13)\*** | — |
| Residual I<sup>2</sup> (%) | 89.3% | 86.7% | 77.1% | 72.5% | 0% |

Values are $\beta$ coefficients with 95% confidence intervals from mixed-effects
meta-regression models. Positive values indicate higher prevalence compared to
the reference category; negative values indicate lower prevalence. Ref:
Reference category for categorical moderators. “—”: Variable not included in the
model or not applicable. MSK: Musculoskeletal problems; PRMS, Playing-related
musculoskeletal symptoms; PRMD, Playing-related musculoskeletal disorders.
\* $p < 0.05$

The prevalence reference period also influenced estimates, studies reporting lifetime prevalence showed significantly lower estimates than those using shorter periods such as 12-month or point prevalence (Model 4: β = –1.03, p = 0.024). Daily practice hours emerged as significant only in the restricted musculoskeletal model (Model 5), where greater practice time was associated with lower prevalence (β = –0.83, p < 0.0001), after adjusting for other moderators. Age group effects were inconsistent, though in Model 5, middle-aged and young adult guitarists had lower prevalence than children, whereas mixed-age samples showed higher prevalence. Residual heterogeneity decreased progressively across models, from 90.7% in Model 1 to 0% in Model 5, indicating that the combined effect of guitar type, age group, prevalence reference period, and practice hours largely explained the between-study variability in musculoskeletal-only analyses.

Sensitivity analyses supported the robustness of these findings. Restricting analyses to studies with clearly specified guitar type markedly reduced heterogeneity, in some cases to zero. Limiting analyses to children or to studies with clinically confirmed PRMDs also reduced heterogeneity, whereas analyses restricted to adolescents or adults retained high heterogeneity. Excluding small studies (<30 participants) did not materially change pooled prevalence estimates. Full numerical results of sensitivity analyses are presented in S2 Table.

### Publication bias

Visual inspection of funnel plots did not suggest substantial asymmetry for the meta-analyses (Fig 6). Egger’s regression test was not statistically significant for the pooled musculoskeletal outcomes (p = 0.7184), suggesting no evidence of small-study effects. However, given the small number of studies in the neurological category (n = 3) and the moderate number in some subgroup analyses, the power to detect publication bias was limited for PRND prevalence. Therefore, the absence of statistical evidence should be interpreted cautiously.

**Fig 6.**
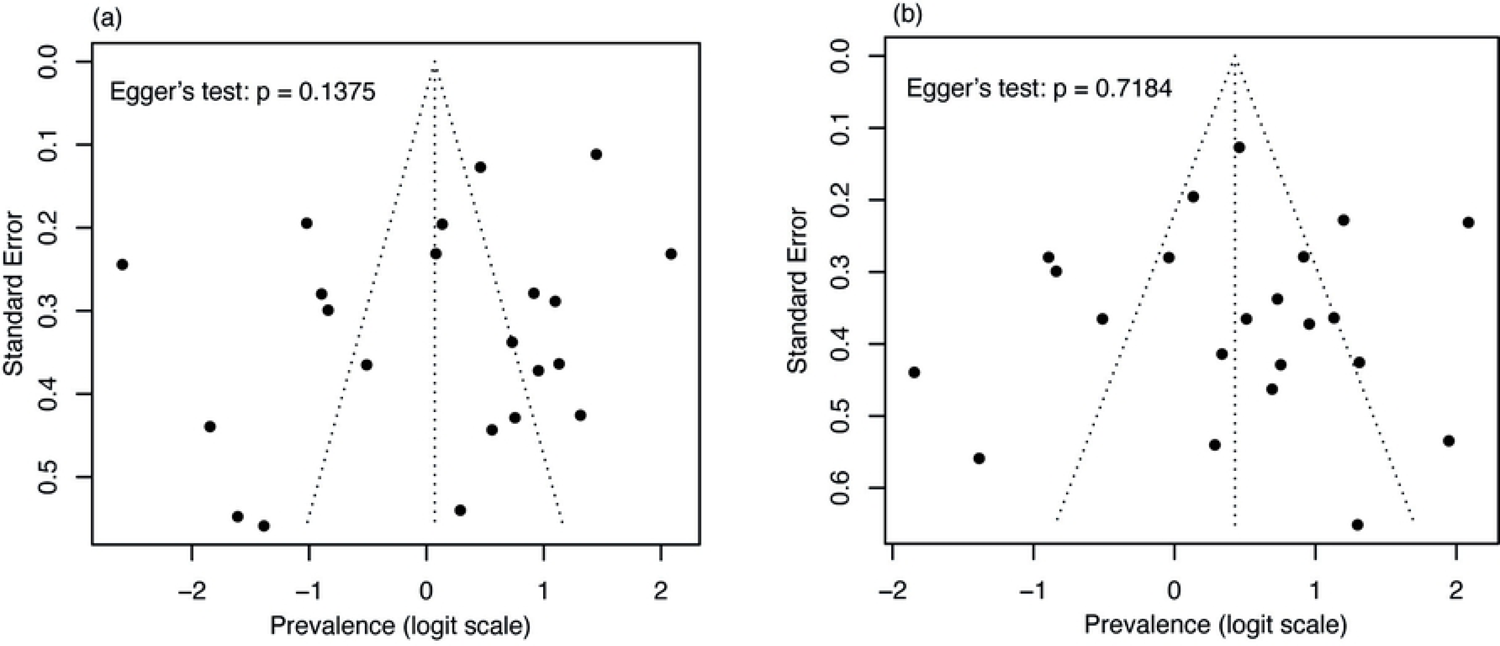
Funnel plots for publication bias assessment. (a) Funnel plot for all playing-related problems (PRP). (b) Funnel plot for musculoskeletal problems (combined PRMD and PRMS). Each dot represents an individual study plotted according to its prevalence estimate (x-axis) and standard error (y-axis). The vertical dashed line indicates the pooled prevalence from the random-effects meta-analysis, and the diagonal dashed lines indicate the expected 95% confidence in the absence of small-study effects.

### Secondary outcomes

Of the 22 studies included in this review, 12 reported at least one secondary analysis exploring potential correlates or modifiers of PRP prevalence (S3 Table). The most frequently examined factors were sex, age group, and practice-related habits. Associations with sex varied by PRP subtype: for focal dystonia, female guitarists had a lower prevalence compared to males in two studies; for PRMS and PRMD, one study reported a lower prevalence in females, one found no significant difference, and one observed a higher prevalence. Older age was more consistently associated with higher prevalence in most studies assessing this variable, except for one with non-significant findings.

Hours of practice per week showed inconsistent associations: most studies found no significant correlation, while one reported a higher prevalence among those practicing 8–14 hours weekly. Warm-up routines and breaks during practice emerged as potential protective factors in some analyses, with significantly lower PRP prevalence among those engaging in these behaviors.

Other less frequently assessed variables included music performance anxiety (MPA), playing position, and engagement in physical activity, with mixed results. Seated playing position was associated with higher prevalence in one study, while another found no significant difference. Physical activity was linked to lower prevalence in two studies, whereas two others found no association. Higher MPA scores correlated with greater symptom burden in one study. The heterogeneity in definitions, measurement tools, and reporting precluded quantitative pooling of these associations.

### Quality of the evidence

All included studies were cross-sectional and most had moderate to severe methodological limitations. Common issues included non-probabilistic sampling, self-selection of participants, inconsistent or vague case definitions, and reliance on self-report without clinical confirmation. Many studies did not report guitar type, playing level, or training background. According to GRADE criteria, the certainty of the evidence for all prevalence estimates was judged to be low, due to risk of bias, indirectness, and inconsistency.

## Discussion

This systematic review and meta-analysis provide the first pooled estimates of playing-related problems (PRP) exclusively in guitarists, revealing that approximately half of this population experience symptoms severe enough to interfere with their ability to perform at their usual level.

Our findings establish a consolidated prevalence of PRPs specifically in guitarists. Previous systematic reviews have offered valuable perspectives on musician health, consistently confirming the high prevalence of PRMDs and the methodological heterogeneity within this research field. For example, the work of Fu and Loo identified that string and keyboard instrument students had the highest probabilities of PRMD [32], while Rotter et al. lamented the lack of differentiation by instrument, which makes it difficult to draw specific conclusions [10]. A systematic mapping review further called explicitly for instrument-specific subgroup analyses and highlighted substantial evidence gaps across instrument families [33]. However, a critical and recurrent limitation in these reviews is the tendency to group instruments with fundamentally different biomechanical demands. This limitation is particularly evident in the classification of guitarists. Notably, no previous systematic review has focused its analysis exclusively on this population. Influential reviews such as Kochem and Silva, which addressed “string players,” focused their analysis exclusively on bowed string instrumentalists (violins, violas, cellos, and double basses), omitting guitarists and other plucked-string players [11]. Other works, such as Rodríguez-Gude et al., identified individual studies on guitarists but presented main conclusions grouped by instrumental families (e.g., upper vs lower strings) and underscored the small number of articles per instrument as a limitation [1]. Grouping guitarists with violinists under a generic “string instrument” category obscures posture, asymmetric loading, and hand-specific kinematics (fretting vs plucking/strumming) that differ fundamentally from bowing.

Even in reviews on specific neurological disorders such as musician’s focal dystonia, where guitarists are recognized as one of the most affected groups along with pianists and violinists, the analysis is rarely broken down to explore the unique risk factors for the guitar [27]. This study directly addresses the gap identified in the literature, providing the first consolidated evidence on the prevalence of PRPs exclusively in guitarists. Based on the evidence found and the results of our meta-analysis—particularly how the sensitivity analysis substantially reduced heterogeneity (from I²=96% to 0%) by focusing on more homogeneous study populations—we suggest the need to move away from broad instrumental groupings. Instead, we recommend that researchers provide specific analyses in their reports with detailed descriptions of the guitar type, age group, experience, and years of practice. This more detailed approach could allow for the detection of risk profiles and the most vulnerable anatomical locations that are unique to guitarists. Consequently, tailored prevention and rehabilitation strategies could be designed and informed for each musician—an objective that, as the previous literature demonstrates, is difficult to achieve when generalized conclusions are applied to instrumentalists with such biomechanical demands.

A fundamental finding of this review is not only the proportion of PRP identified but also the significant methodological limitations inherent in the primary studies, which requires a cautious interpretation of our meta-analysis results. Although the risk of bias assessment revealed serious concerns in most studies, particularly regarding sampling, participant selection, and low response rates. The primary meta-analysis of proportions that included all studies showed extremely high heterogeneity (I² = 96%). This variability appears to stem from differences in outcome definitions and measurement such as PRMD versus PRMS and variation in study populations, including instrument type (classical, bass or mixed acoustic/electric) and age group. In sensitivity analyses, heterogeneity was eliminated or markedly reduced when restricting to highly specific subgroups: studies of children (I² = 0%), and studies that specified instrument as bass or classical guitar (I² = 0% for each), with a substantial reduction for mixed acoustic/electric guitar (I² = 34%). These findings indicate that the high heterogeneity reflects aggregation across disparate populations, instruments, and outcome definitions rather than random variation.

Secondary outcomes reported by 12 of the 22 included studies provide additional insight into potential risk or protective factors for PRP (S3 Table). Associations with sex varied by PRP subtype, in the focal dystonia, female guitarists had a lower prevalence compared to males in two studies [22,27]; for PRMS and PRMD, one study reported a lower prevalence in females [17], one found no significant difference [20], and one observed a higher prevalence [26]. Older age was more consistently associated with higher prevalence in most studies assessing this variable [17,22,25–27], except for two with non-significant findings [20,24]. Notably, this pattern contrasts with previous systematic reviews encompassing multiple instruments (e.g., orchestral settings), where female sex has generally been associated with a higher risk of musculoskeletal disorders. The divergence observed here suggests that sex-related risk may be instrument- and disorder-specific. These findings reinforce the importance of conducting instrument-specific and condition-specific analyses, as aggregating disparate instruments and disorder types may obscure meaningful differences in risk profiles driven by biomechanical demands. Hours of practice per week showed inconsistent associations, most studies found no significant association [4,17,18,20,26], while one reported a higher prevalence among those practicing 8–14 hours weekly [30]. Warm-up routines and breaks during practice emerged as potential protective factors in some analyses, with significantly lower PRP prevalence among those engaging in these behaviors [4,20,23,25]. Other less frequently assessed variables included MPA [5,24], playing position, and engagement in physical activity, with mixed results. Seated playing position was associated with higher prevalence in one study [25], while another found no significant difference [20]. Physical activity was associated with lower prevalence in two studies [25,30], whereas two others reported no significant association [20,23]. Higher MPA scores correlated with greater symptom burden in one study [25]. The heterogeneity in definitions, measurement tools, and reporting precluded quantitative pooling of these associations, but they highlight modifiable factors that could be targeted in prevention programs.

## Conclusions

This systematic review and meta-analysis consolidate, for the first time, the prevalence of PRPs in guitarists, demonstrating that these disorders affect a substantial proportion of musicians and can significantly compromise performance capacity. By disentangling problem subtypes and instrument categories, our findings reveal that biomechanical demands and disorder classification are critical determinants of risk and heterogeneity. While methodological limitations in the primary literature require cautious interpretation, the consistent patterns observed—together with suggestive evidence on modifiable risk factors such as warm-ups, breaks, and physical activity—offer a foundation for prevention strategies tailored to the unique demands of guitar playing. Addressing these health issues through targeted interventions, early screening, and ergonomic education has the potential to reduce injury burden, improve career longevity, and enhance the well-being of guitarists. This study underscores the urgent need for standardized definitions, rigorous longitudinal research, and more granular reporting to advance evidence-based practice in performing arts medicine.

## Data Availability

All relevant data are within the manuscript and its Supporting Information files.

## Acknowledgments

We are grateful to Mstr. Jorge Lay-Herrera, Mstr. Víctor M García-Ramírez and Mstr. José L Segura-Maldonado for their expert insights into the differences in various guitar types of construction, classification, and performance techniques. This contribution was crucial for understanding the distinct biomechanical demands of each instrument and for contextualizing the findings of this study.

---

**S1 File.** PRISMA checklist.

**S2 File.** Detailed search strategy.

**S1 Fig. Risk of bias summary for each included study**. This figure summarizes the risk of bias assessment for each study included in the systematic review. Each row corresponds to a specific study, while each column represents a bias domain. The plot is color-coded as follows: a green circle indicates a low risk of bias, a yellow circle indicates an unclear risk of bias, and a red circle indicates a high risk of bias for that specific domain.

**S1 Table. Characteristics of included studies.** Study-level variables for PRP in guitarists: study ID, country, population, N, guitar type, PRP subtype (PRMS/PRMD/PRND), case definition, prevalence with recall period, anatomical site, and secondary outcomes; denominators flagged when conditional on PRP.

**S2 Table. Sensitivity analyses for pooled prevalence of PRP**. Random-effects scenarios: specified guitar type; age group; clinically confirmed PRMD/PRND; excluding n<30; stratified by recall period (point, 3-month, 12-month, lifetime).

**S3 Table. Summary of secondary outcomes across included studies**. Secondary outcomes (practice time, warm-up, sex, multi-instrumentalism, posture/technique, breaks, etc.) with effect estimates (OR/PR/RR), 95% CI/p-values, adjustment, and brief notes; not pooled.

